# Using interpretable random forest to determine observational-data-based cutoff points in medical measurements

**DOI:** 10.1101/2025.07.10.25331301

**Authors:** Wentian Li

**Author notes:** The Feinstein Institutes for Medical Research, Northwell Health, Manhasset, NY, USA.

## Abstract

Regular checkups are crucial in health management of a general population. Part of a checkup includes monitoring substances in blood, in urine, as well as physical measurements related to health. It is often taken for granted that normal ranges, specified by cutoff points, of these measurements are rigorously determined by scientific evidence. In reality, there is a degree of uncertainty due to possible change of the definition of being healthy, small sample sizes in intervention trials, and gender, ethnic group, and individual differences. One conceptual difficulty in setting a cutoff point is that generally there is no evidence for a threshold effect – quantum jump of risk for detrimental medical outcome once the measurement level crosses the cutoff point – for almost all physical checkup measurements. A solution in unambiguously determining a cutoff point at the population level, even though the risk changes continuously, is by minimizing the sum of two false classification rates (false positive and false negative rates), as one false rate will increase whereas another decrease with the change of the measurement level. Another problem in cutoff determination is that cutoff of a measurement may depend on the context of other variables such as gender or other components in blood. We use interpretable random forests (IRF) to tackle this latter problem. The output from an IRF is a set of node split rules involving one or multiple variables. Each rule is a possible cutoff point, and a composite rule provides a cutoff point conditional on other variables. We use the National Health and Nutrition Examination Survey (NHANES) data to illustrate this approach, as well as to show that the observational-data-determined cutoff point depends on the data used, the medical outcome chosen, and the cost or objection function employed.

## Introduction

A patient’s annual physical examination or checkup (AMA Council on Scientific Affairs, 1983; Frame, 1995; Chang et al., 2002; Boulware et al., 2007) consists of discussion of any unusual symptoms with the patient’s family doctor, checking vital signs, plus blood tests (Carmalt et al., 1970; Leurquin et al., 1995; Watson et al., 2017) and urinalysis (Simerville et al., 2005; Zhang et al., 2022), etc. If vital signs, chemical properties in serum, urine, or any medical measurements are out of the “normal range”, it is a concern for the patient’s health, and further actions need to be taken. One might think that normal range of any measurement level is just determined by a population histogram, which might follow a normal distribution; and the normal range is determined by a *z*-standard-deviation from the mean. However, it is not the case: being an outlier does not automatically imply a health hazard. For example, exceptionally tall or short people can still have a normal life span. The justification for what is considered as normal and what is not must be explained explicitly.

Needless to say, it would be ideal to derive a normal range or cutoff values from first principles in biochemistry, biophysics, or physiology. Blood pressure is decided by three forces in the arteries: elastic, kinetic and gravitational (Magder, 2018). However, it is rarely possible to obtain clinically relevant results from the first principle, such as a normal range of blood pressure. The next best option is to carry out medical intervention experiments. For example, in a stepped care program, patients with diastolic blood pressure (DBP) above 90 mmHg (and for those within the range of 90-104 mmHg) treated with anti-hypertensive therapy had significant (at the level of *p <* 0.01) lower 5-year mortality rate than the referred care group without treatment (Hypertension Detection and Follow-up Program Cooperative Group, 1979, 1982). A similar trial in treating people with systolic blood pressure (SBP) over 130 mmHg without diabetes indicates that lowering SBP to around 120 decreased the rate of one year detrimental cardiovascular events from 2.19% to 1.65% (*p <* 0.001) (SPRINT Research Group, 2015). These types of interventional trials would be a direct proof that DBP higher than 90 mmHm or SBP higher than 120mmHm is not good for health.

Without an intervention program, one may settle on population data. If the probability of mortality or morbidities as a function of the medical measurement is U-shaped, the span of the bottom region can be used to define the normal range. If it is J-shaped or L-shaped, a high-end or low-end cutoff value can be determined. For example, the relationship between body mass index (BMI) in 50-year-old men and mortality followed for 30 years, is U-shaped (Troiano et al., 1996). The bottom of the U-shaped curve led to a normal range of 23-28 kg/m^2^ for BMI (Troiano et al., 1996).

The normal range and cutoff values for medical measurements are usually given “as it is” without more explanations. Many of these cutoff values are resulted from committee reports. However, experts may not always agree with each others. One may see these sentences in some of the committee reports, such as: “the method used to established BMI cut-off points have been largely arbitrary. In essence, it has been based upon visual inspection of the relationship between BMI and mortality … studies in this area have usually suffered from certain methodological drawbacks” (page 313 of (WHO Expert Committee, 1995)); “the cut-point values used to define (vitamin-D) deficiency · · · have not been established systematically using data from studies of good quality. Nor have values to be used for such determinations been agreed upon by consensus within the scientific community” (page 8-1 of (Ross et al., 2010)).

One common misunderstanding of the cutoff value is that there is always a threshold effect, i.e., a quantum jump of risk for mortality or morbidities when the medical measurement level exceeds the cutoff value (threshold effect in neuron firing, on the other hand, is real and essential (Hodgkin and Huxley, 1952; Gerstner et al., 2014)). Admittedly, if a diagnostic biomarker is the sole cause of a disease, or is a unique waste product of one, but not other, disease, its non-zero level should signal the presence of that disease. Genetic diseases or viral diseases often belong to this category.

However, for chronic and complex diseases, threshold effect is either rare or nonexistent. In these situations, the chance for deleterious medical consequences will most likely increase continuously. Individuals with, e.g. higher diastolic blood pressure or BMI or lower vitamin-D than the normal cutoff level can still experience no health problems (false positives), and those with lower DBP or BMI or higher vitamin-D level can still have harmful medical outcomes (false negatives). In this situation, a cutoff (or two cutoffs which bracket the normal range) is merely a trade-off or compromise between the false positive rate and the false negative rate at the population level (page 14 of (WHO Expert Committee, 1995)).

The above understanding, that most expert-provided cutoff values are optimal points that minimize false prediction rate (some combination of false positives and false negatives), provides a mathematical framework for precisely defining a cutoff value, when such value is determined by observational data. Suppose *x*(*t*_1_) is a medical measurement at time (or age) *t*_1_, and *y*(*t*_2_) is a binary medical outcome at time (or age) *t*_2_(*> t*_1_), and suppose the risk to have harmful medical outcome (*y* = 1) increases with the measure *x*, then the population level probability for false positive could be *Prob*(*x*(*t*_1_) *> X*|*y*(*t*_2_) = 0) (false-positive rate (FPR), equal to 1 minus specificity) and false negative can be *Prob*(*x*(*t*_1_) *> X*|*y*(*t*_2_) = 1)) (false negative rate, equal to 1 minus sensitivity) where *X* is an arbitrary cutoff value. A simple combination of the two types of errors is the average of the two; then, the best cutoff is defined as the *X* value that minimizes the combined error, e.g.

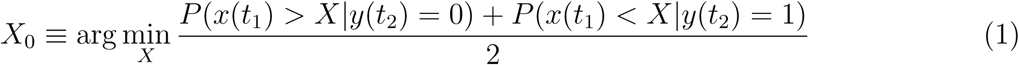

There can be other variations of the above formula. For example, (i) the two types of error can be weighted by *w*_0_ and *w*_1_, instead of 1/2 each, and the weight might be the population prevalence (*w*_0_ = *P* (*y* = 0)*, w*_1_ = *P* (*y* = 1)). (ii) the single cutoff point *X*_0_ can be expanded to an interval (*X*_0_*_l_, X*_0_*_r_*) where the combined errors are among the lowest but not the absolute low (note that this mechanism to define an range is different from the situation where the risk itself is U-shaped). (iii) switching the event and condition in the definition of FPR becomes the false discover rate (FDR) and doing that in the definition of FNR leads to false omission rate (FOR). Then one may determine the cutoff by minimizing the mean of FDR and FOR. (iv) instead of minimizing a function (e.g., combined error), one may also maximize a function. A trivial example on this is that we may maximize the sum of sensitivity and specificity, which is the same idea behind the Youden index (Youden, 1950).

As these mathematical definitions of cutoff points are intrinsically related to a classification problem (and the classification errors), similar classification methods in machine learning (ML) or artificial intelligence (AI) can also be considered. In fact, the classification trees (Breiman et al., 1984) (or recursive partitioning (Zhang and Singer, 2010), or decision trees (Quinlan, 1986)) uses greedy algorithm to scan all possible split points that either minimize some quantities, e.g., (weighted sum of) Gini impurity, entropy, or maximize some quantities, e.g., distance between members in different groups after split, including the Hellinger distance (Cieslak et al., 2012). Random forests (Breiman, 2001), as an extension of the classification and regression trees, also deal with the cutoff selection issue in individual trees. These computer-science-field developed approaches are parallel to the similar developments in the field of statistics.

With population-based cutoff definitions like Eq.(1), many potentially confusing questions can now be addressed clearly, and misunderstandings can be avoided. First of all, one needs to know which sample population or subset of patients are used to derive the cutoff. Different gender, age, ethnicity groups may have different cutoff points. Secondly, which medical outcome *y* is used may impact the result: *y* can be mortality (and mortality after up to certain number of years), a particular disease, or a basket of morbidities (combined with certain weighting choices). Thirdly, information should be given on which cost function (or loss function, error function, to be minimized) or objective function (or utility function, payoff function, fitness function (in evolutionary biology), to be maximized) is used in the determination of a cutoff value. Using different cost or objective functions could change the cutoff value.

Extension to the first point above, cutoff values could also be different between smokers and non-smokers, people with different genetic predispositions, people with different risk factor status. These represent different subsets in a population. Ultimately, there should be a personalized cutoff value specific to anyone with a particular condition. This last point, that a particular subset with specific status for other risk factors should have its own specific cutoff value, can be addressed by a machine learning approach. In this context, the cutoff determination problem is no longer a relationship between one univariate *x* and an outcome *y*, but between multiple variates (*x*_1_*, x*_2_, · · · *x*) and *y*.

In this work, we propose the use of interpretable random forest (IRF) (Aria et al., 2021) to find cutoff points for appropriate subsets; in other words, the cutoff value for one variable that are conditional on other factors. There are several considerations behind this approach. First, ML or AI techniques (where random forest is an example) have an important advantage over traditional statistical methods in that the default setting is multiple-variate. Therefore, a result from ML in general, and random forest in particular, is already in a context of other variables. Second, to extract interpretable results from a ML method, it is better to use an “inherently interpretable ML model” (Du et al., 2020; Rai, 2020) than a “black-box model” such as artificial neural networks and deep learning. Random forest is based on decision trees, with the latter having a high level of interpretability. From decision trees and random forest, although the interpretability is greatly reduced (see Fig. 9 of (Hassija, 2023)), the two are still closely related that a solution is possible. Third, the node splitting in decision trees and random forest is in fact a cutoff determination process. This would make them an ideal fit to our goal.

The paper is organized as follows: the first level consists of Introduction, Results, Discussion, Data and Programs. Then we have several sub-sections within the Result section. The first Result subsection, using a simple theoretical model to formulate our cutoff determination process, is mostly presented in the Appendix A.2, as it is more an overview than a new result. Then, in the next Result subsection, we use the National Health and Nutrition Examination Survey (NHANES) data to show that the cutoff determination could depend on which medical outcome is used. The next subsection discusses several objective functions in the context of decision trees. Then in two main subsections, the cutoff determination by the interpretable random forests is illustrated.

## Results

### A theoretical toy model for the determination of cutoff point

We use a simple model to illustrate the basic concepts and conclusions concerning databased determination of cutoff point by minimizing the mean of two types of classification rates. The model is a surrogate of real data, with an advantage that many results might be obtained analytically.

Our toy model assumes the medical measurement *x* for the healthy group (*y* = 0) and for the diseased group (*y* = 1) both follows a normal distribution, but with different means (though same variance). Any cutoff point *X* will cut both distributions into two parts, a part below *X* and another part larger than *X*. With two parts in two distributions (healthy and diseased), there are four parts altogether (see Appendix A.1). Two parts of these two represent incorrect or false classifications, and the other two correct ones. Therefore, the mean of the two false classification parts as a function of cutoff point *X* can be plotted directly.

From this model, we can understand that the risk for disease is a gradual and continuous function of the cutoff point *X*. We can also see that if the two classification rates are weighted, the minimal position for the cost function is also changed.

For our specific model, besides the minimal position for the cost function, there are also two other ways to determine the cutoff point. One is the value of *x* where the two distributions intersect. Another is the value of *x* where the cumulative distributions of the two have the largest difference. All detailed derivation of these results can be found in Appendix A.2.

### Cutoff value determination may depend on the medical outcome used

The 2017-2018 NHANES data (adults who are 20 years old or older) is used to show that the optimal BMI value depends on which medical outcome is used: whether one wants to minimize the false rate for classifying diabetes or classifying heart problems. Fig. 1 shows the cost function (FPR + FNR)/2 as a function of the cutoff used with diabetes (black) or any one of the five heart conditions (blue) as *y*. The BMI values around 29.9 ∼ 31.2 achieve the minimum cost function for classifying diabetes, whereas the BMI values around 27.5 ∼ 27.8 correspond to the best result for classifying heart diseases. This result indicates that cutoff value determination for BMI, even when a cost function has been chosen, depends on which medical outcome is used as the targeting outcome.

**Figure 1:**
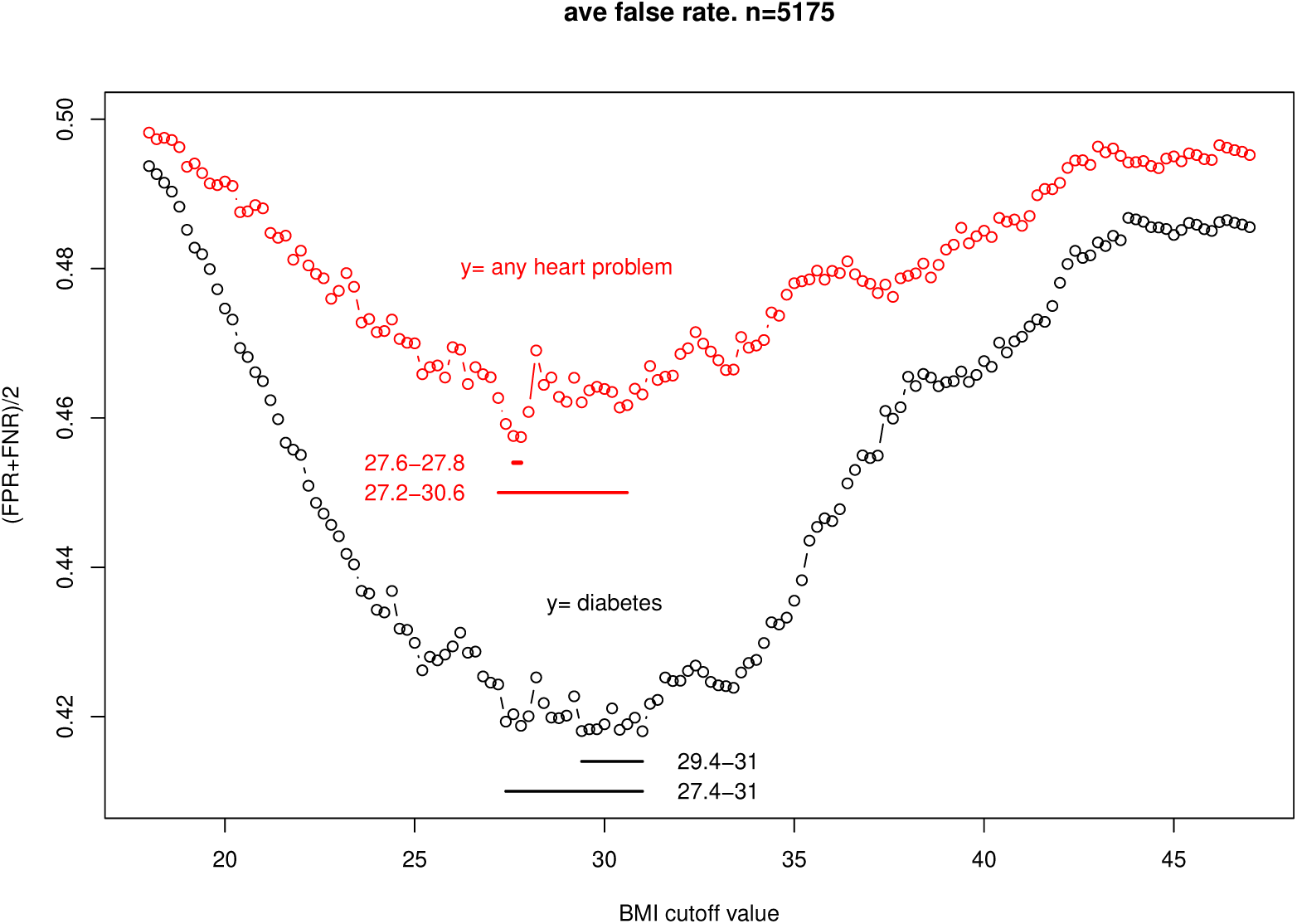
The mean of two false rates, false positive rate and false negative rate ( (FPR+FNR)/2), as a function of BMI cutoff value: (black) diabetes as the outcome, (red) any heart problem as the outcome.

If the false positive rate is given more weight (*w*_0_ = 1 − *P* (+), *w*_1_ = *P* (+), where *P* (+) is the prevalence of the disease: P(diabetes)=0.19 and P(heart disease)=0.12 for this dataset), the minimum of the cost function is reached at the maximum possible BMI value (plot not shown). The reason for this, as already mentioned in the last subsection, is that the cost function is dominated by the FPR, which will decrease with increasing cutoff value; there is no balancing act from the FNR within the range of observed BMI values, not only because of its small contribution, but also because the balancing point may be beyond the upper limit for possible BMI values.

Due to limitation of any particular dataset, there are also noise and uncertainty is the cutoff determination. One way to solve this problem is by a meta analysis pooling data from different studies. Another solution is to relax the requirement of absolute minimization, by combining all *x*’s that have *y < y*_0_, instead of *y* = *y_min_*. From that consideration, the whole BMI region 27 ∼ 31 has relatively low false rates ((FPR+FNR)/2) in classifying both diabetes and heart problems.

### Fisher’s test *p*-value as an alternative measure of sample separation performance by the cutoff

In the last subsection, a combination of false classification rates FPR and FNR is used as the cost function whose minimal point is searched. Besides false classification rates, there are other measures that characterize the success or failure in partitioning the samples into two groups. In this subsection, we investigate other possibilities and their potential impact on cutoff determination. The medical outcome is the (doctor’s diagnosis of) diabetes, and the independent variable *x* is BMI.

Any cutoff point will partition the data into 4 groups, conveniently forming a 2-by-2 table (see Appendix A.1). The samples in the matrix diagonal elements (TN and TP, see Appendix A.1 are correctly classified, whereas those in the off-diagonal (FN and FP, see Appendix A.1) are falsely classified (errors). The FPR and FNR are FP and FN normalized by the actual numbers of healthy and diseased samples. Other measures to characterize the 2-by-2 count table include (diagnostic) odds-ratio (TN · TP)/ (FN · FP), relative risk, and Fisher’s test *p*-value for the 2-by-2 table, which is a measure of statistical significance. The test *p*-value actually behaves similarly as (FPR+FNR)/2 and will be the measure we use here.

Fig. 2 shows the log_10_(*p*-value) as the function of cutoff of BMI. The lower the *p*-value, the more unlikely that differences between two subsets of samples partitioned by a cutoff is a chance event. We therefore search for BMI that minimizes the *p*-value. The lowest *p*-value is reached at BMI=23.5, and the bracket (22.5-25) is the lowest region. By scanning the Fig. 2, the region (22-31) tends to contain lower *p*-values. As a comparison, we redraw part of Fig. 1, with diabetes as the medical outcome, that shows (FPR+FNR)/2 curve, on Fig. 2. The lower region for the average error rate is (27.5-31).

**Figure 2:**
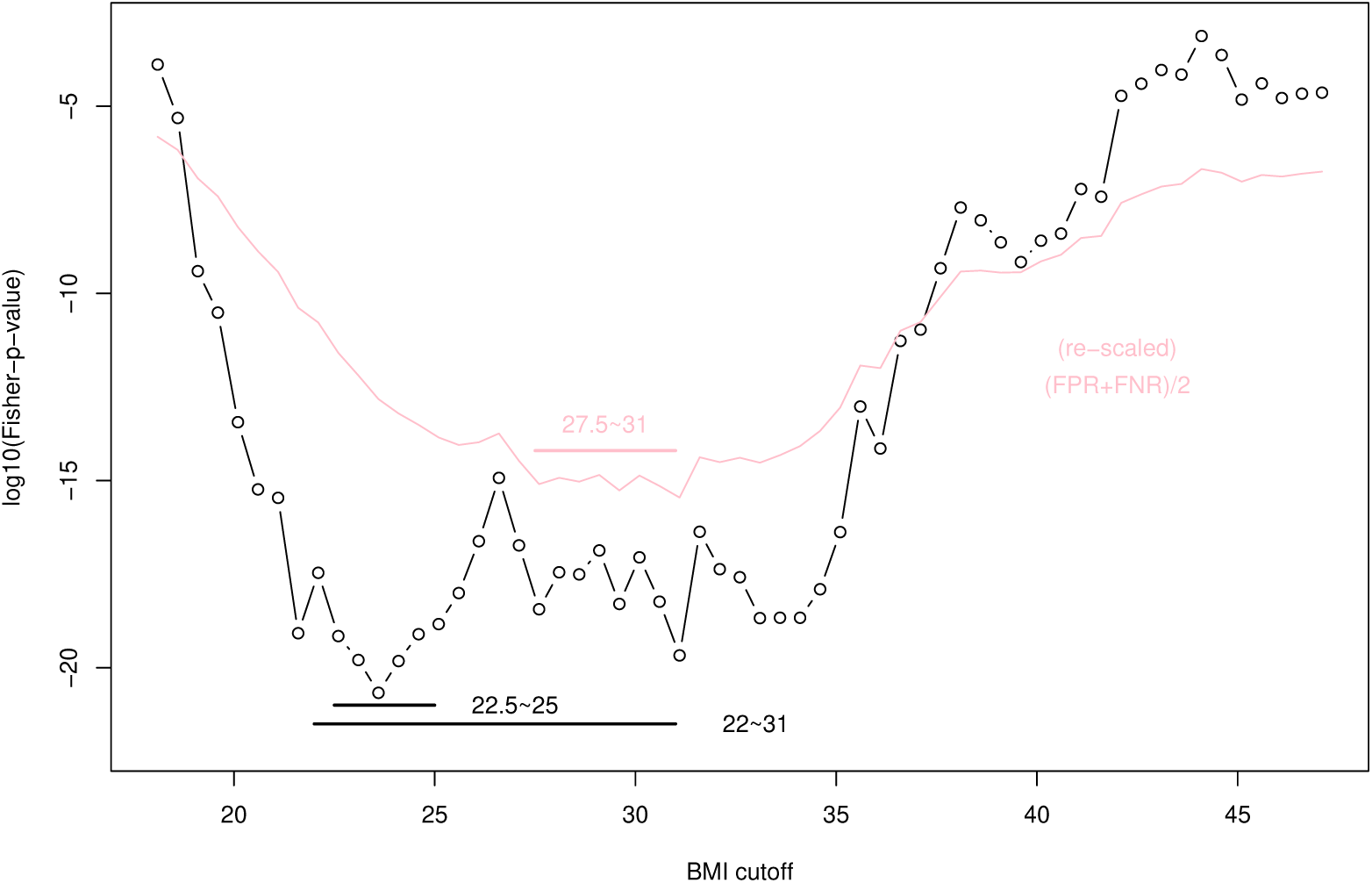
Fisher test *p*-value (log_10_) as a function of the BMI cutoff value, with diabetes as the outcome variable. The average false rate (from Fig. 1, black line) is reproduced here (in red) as a comparison (the *y* scale is chosen arbitrarily).

Fig. 2 shows that even with the identical data, identical medical outcome *y*, changing from (FPR+FNR)/2 to *p*-value (two different measurements used to characterize the separation of the two groups, *BMI < T* and *BMI > T*), the cutoff point determined can be different. This is the case despite the general similarity between (FPR+FNR)/2 and Fisher’s *p*-value (the two are both lower in the middle area of BMI).

### Cutoff determination in decision trees depends on the choice of cost or objective function

Many methods in machine learning need a cutoff determination in their implementation, including decision or classification trees and random forests. In a split of node into children nodes for a decision tree, it is required that the (weighted sum of) impurities of the children nodes is lower than that of the root node. A popular choice for the measure of impurities is the Gini index, which is simply equal to 2P(+)P(−) in the root node. The weighted Gini index in the children nodes is 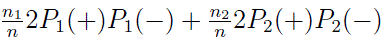, where *n*_1_ is the number of samples in the first node (e.g. for *x < X*), *n*_2_ is that in the second node (e.g. for *x > X*), *n* = *n*_1_ + *n*_2_, and *P_i_*(+), *P_i_*(−) are the proportion of *y* = 1, *y* = 0 samples in node *i* ∈ (1, 2).

The entropy-based split rule requires that the (weighted sum of) entropies in the children nodes is lower than that in the root node. The latter is −*P* (+) log_2_ *P* (+)−*P* (−) log_2_ *P* (−), and the former is 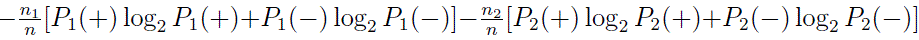. Without the negative signs, entropy will become information, and the split rule requires an increase of information in the children nodes than that in the root node. Note that neither Gini index nor entropy need to know that true negative is the majority in the first node or true positive is the majority in the second node. When the population prevalence *P* (+) is low, *y* = 1 samples may be the minority in both nodes. The key point is whether a node split greatly changes the contrast in these two nodes as compared to the root node.

More recently, a new node split criterion was introduced that is suggested to be insensitive to imbalanced data, using the Hellinger distance (Cieslak and Chawla, 2008; Cieslak et al., 2012). Hellinger distance is an Euclidean distance in the square-root-transformed probability space (Hellinger, 1909). The measure used in node splitting is defined as the following:

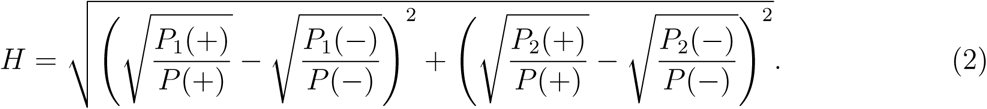

It is the distance between the two (square-root-transformed and relative) distributions for *y* = 0, 1 samples in the children nodes. The “relative” part refers to a normalization of the children node distribution by the distribution in the root node. Imbalanced data is a problem (He and Garcia, 2009) when the population prevalence is low (Khalilia et al., 2011) and when the data is not collected by a case-control design.

Fig. 3 plots the three objective functions: (1) the amount of decrease of Gini impurity (normalized by the maximum possible value), (2) decrease of entropy (normalized by the maximum possible value), and (3) Hellinger distance (divided by 7 to be visually comparable to the other functions), for classifying diabetes, as a function of BMI cutoff point. For Gini index decrease, the maximum is reached at BMI= 33.7 (black line); for entropy decrease, the maximum is reached at BMI=23.7; and for Hellinger distance, the maximum is at BMI=21.7. Despite the different results in cutoff determination using different measures, the overall shape of the objective functions are nevertheless similar. A range of BMI 21.7 ∼ 33.7 represents the plateau region of these curves.

**Figure 3:**
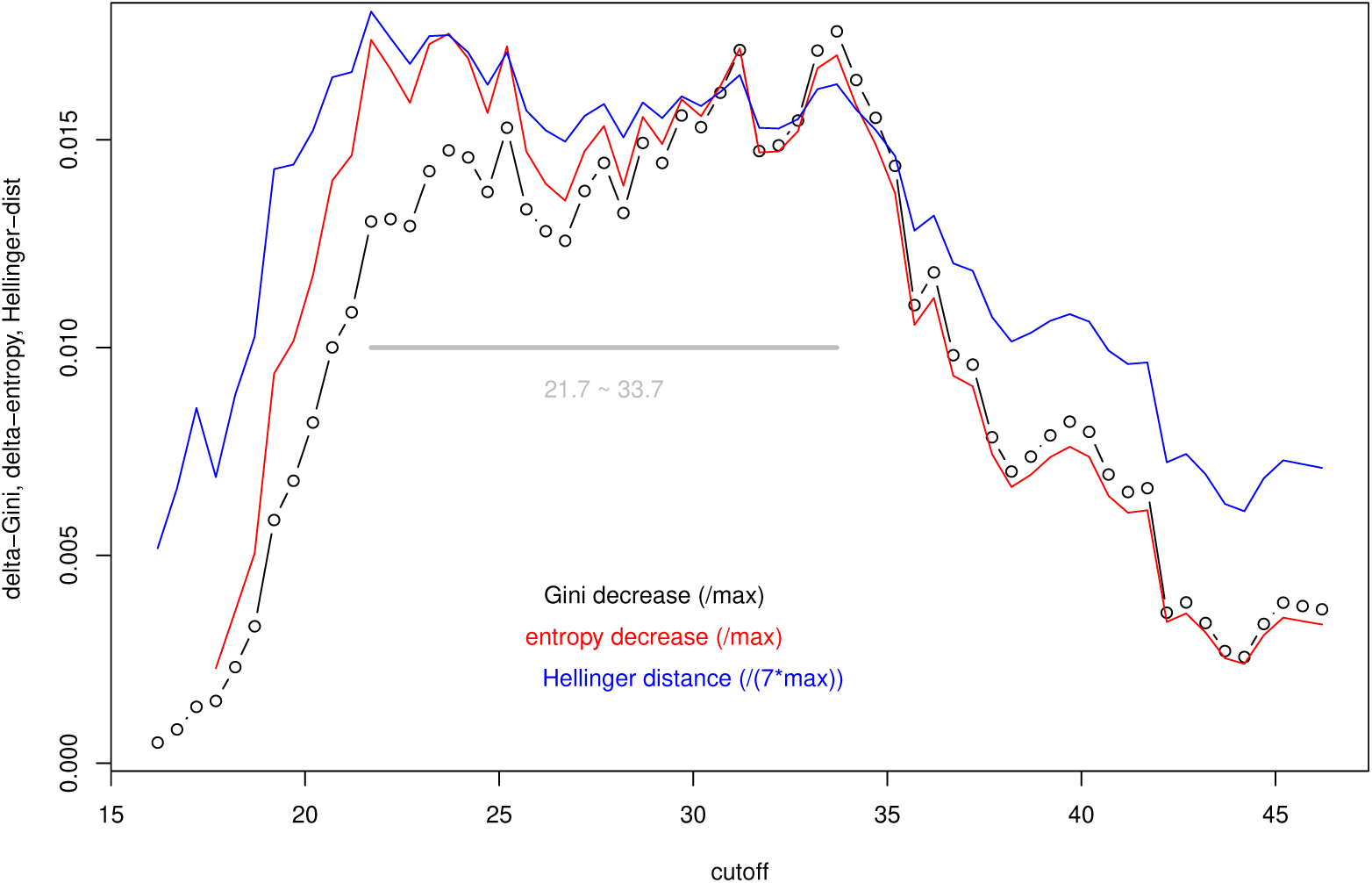
Three objective functions as a function of cutoff value for BMI (diabetes is the outcome variable): decrease of the (normalized) Gini impurity with a partition of samples by the cutoff point (red); decrease of the (normalized) entropy with a partition of the samples by the cutoff (red); Hellinger distance (Eq.(2)) between before and after a partition of samples by the cutoff (blue, scale is chosen arbitrarily to match other curves). The region with high *y* values is marked by a horizontal line.

### Conditional cutoff determination by interpretable random forests

Random forests are ensemble (multiple) decision trees. Each tree fits a subset of the samples, with its own variable selection and cutoff determination. Result from a random forest summarizes those from individual trees. Although random forests lag behind decision trees in their interpretability, the close relationship between the two makes an interpretation of random forests possible. effectively converting seemingly “black-boxes” into “transparent-boxes” or “glass-boxes”.

It is easy to see why the node splitting in decision trees or random forests is different from a single-variable cutoff determination. In the simple case of two-variables trees or forests, *x* is the variable of interest, and *x*_1_ is the co-variate; if the root node is split according to value of *x*_1_, and the child nodes being further split by *x*, the cutoff determination for *x* is carried out conditional on the *x*_1_ variable. Therefore, the determined cutoff point for *x* is within the context of *x*_1_.

Although training results from decision trees are themselves rules, these tend to unstable with respect to minor changes in the data. Ensemble of trees such as random forests, on the other hand, produce more reliable results. There have been many works to extract rules from ensemble of trees (Friedman and Popescu, 2008). We adopt a more recent program, SIRUS (Stable and Interpretable RUle Set for classification), claimed to be more stable than other approaches (Bénard et al., 2021). The main goal of SIRUS is to find more frequently appearing hyperrectangles, then combine and summarize them into simple rules. The SIRUS program pre-discretizes continuous variables into *q*-quantiles (usually *q* = 10). It has mostly one superparameter that can be adjusted to influence the number of rules to be produced. This superparameter is set in such a way that at most 25 rules are printed out (Bénard et al., 2021; Bénard, 2021).

We ran SIRUS with BMI, age, gender as three possible predictors, and diabetes as the medical outcome. Table 1 shows part of the SIRUS generated rules that contain BMI (other rules from the program output could involve only age, only gender, or only age and gender). The SIRUS program has selected three BMI-only cutoff values: 23.9, 30.3, and 35. Even though FPR and FDR are both measures of false positives, they can be sometimes very different, as their denominators are different. For example, in ensemble average, there are 1039 samples with BMI ≥ 35, 743 of them are non-diabetes. The FDR is 743/1039=71.5%, but FPR is 743/4199=17.7% where 4199 is the total number of non-diabetes. Table 1 shows that FDRs are consistently above 70%, FORs are around around 10%-16%, and both (FPR+FNR)/2 and (FDR+FOR)/2 are around 42%-45%.

**Table 1:** Rules produced from SIRUS with BMI, age, gender as independent variables, and diabetes as the dependent variable. Total four rules in the output involve BMI: three are BMI only, and one involves BMI and gender. Each rule partitions the samples into 4 groups (TN, FN, FP, TP) in a 2-by-2 table. *n*_1_=TN+FN is the number of samples in the first node, and *n*_2_=TP+FP is that in the second node. From the 2-by-2 table, we calculate the FNR, FOR, FPR, FDR, (FPR+FNR)/2, (FOR+FDR)/2, odds-ratio, and Fisher test *p*-value (see Appendix A.1 for definitions).

| condition0 | $n_1$ | FNR | FOR | condition1 | $n_2$ | FPR | FDR | $\frac{FPR+FNR}{2}$ | $\frac{FOR+FDR}{2}$ | p-value | OR |
| --- | --- | --- | --- | --- | --- | --- | --- | --- | --- | --- | --- |
| BMI < 23.9 | 1004 | 0.094 | 0.092 | BMI >23.9 | 4171 | 0.783 | 0.788 | 0.438 | 0.44 | 1.4E-20 | 2.67 |
| BMI < 30.3 | 3091 | 0.469 | 0.148 | BMI >30.3 | 2084 | 0.373 | 0.751 | 0.421 | 0.45 | 1.7E-19 | 1.9 |
| BMI < 35 | 4136 | 0.695 | 0.164 | BMI >35 | 1039 | 0.177 | 0.715 | 0.436 | 0.44 | 1.3E-17 | 2.0 |
| else | 4231 | 0.733 | 0.169 | BMI ≥30.3<br>and male | 944 | 0.162 | 0.721 | 0.447 | 0.445 | 6.8E-14 | 1.9 |

The Fisher’s test *p*-values and odds-ratio of the 2-by-2 matrices formed by TN, FP, FN, TP, are also shown. The *p*-values are all very low (very significant), and the odds-ratios are all around or larger than 2. These very good statistical results are in striking contrast with the poor classification performances (40% plus errors). Part of the reason is the large sample size used here, which will make even a small signal unlikely to be produced by chance event (thus statistically more significant).

It is interesting that determined cutoff values from these random forest results are comparable to the single decision tree result in Fig. 3, as both 35 and 23.9 are close to peak positions, and 30.3 is well bracketed in the high plateau.

Besides single-variable (BMI only) rules, SIRUS also obtains a new rule containing BMI and gender (last row of Table 1): for male gender with BMI ≥ 30.3, there is a higher chance to develop diabetes than samples do not satisfy this condition. Although the overall error rate from this rule (either (FPR+FNR)/2 or (FDR+FOR)/2) does not improve upon BMI-only rules, it does, however, show that the combination of high BMI and male gender could lead to a higher risk for diabetes.

To check if this new rule obtained from SIRUS makes sense, we plot the averaged error (FPR+FNR)/2 as a function of BMI cutoff in male and female samples separately (Fig. 4). Fig. 4 shows that the cutoff for female samples would be chosen at BMI=27.4 as it minimizes the mean error, whereas the cutoff for male samples would be BMI=31. Actually Fig. 4 shows that besides the global minimum, female samples exhibit another local minimum around BMI=33. On the other hand, for male samples, there is only one minimal region around 29.8∼31.2. These three minimal are reflected by the three cutoff values obtained from SIRUS in Table 1. The last line in Table 1 is also consistent with the male only cutoff seen in Fig. 4.

**Figure 4:**
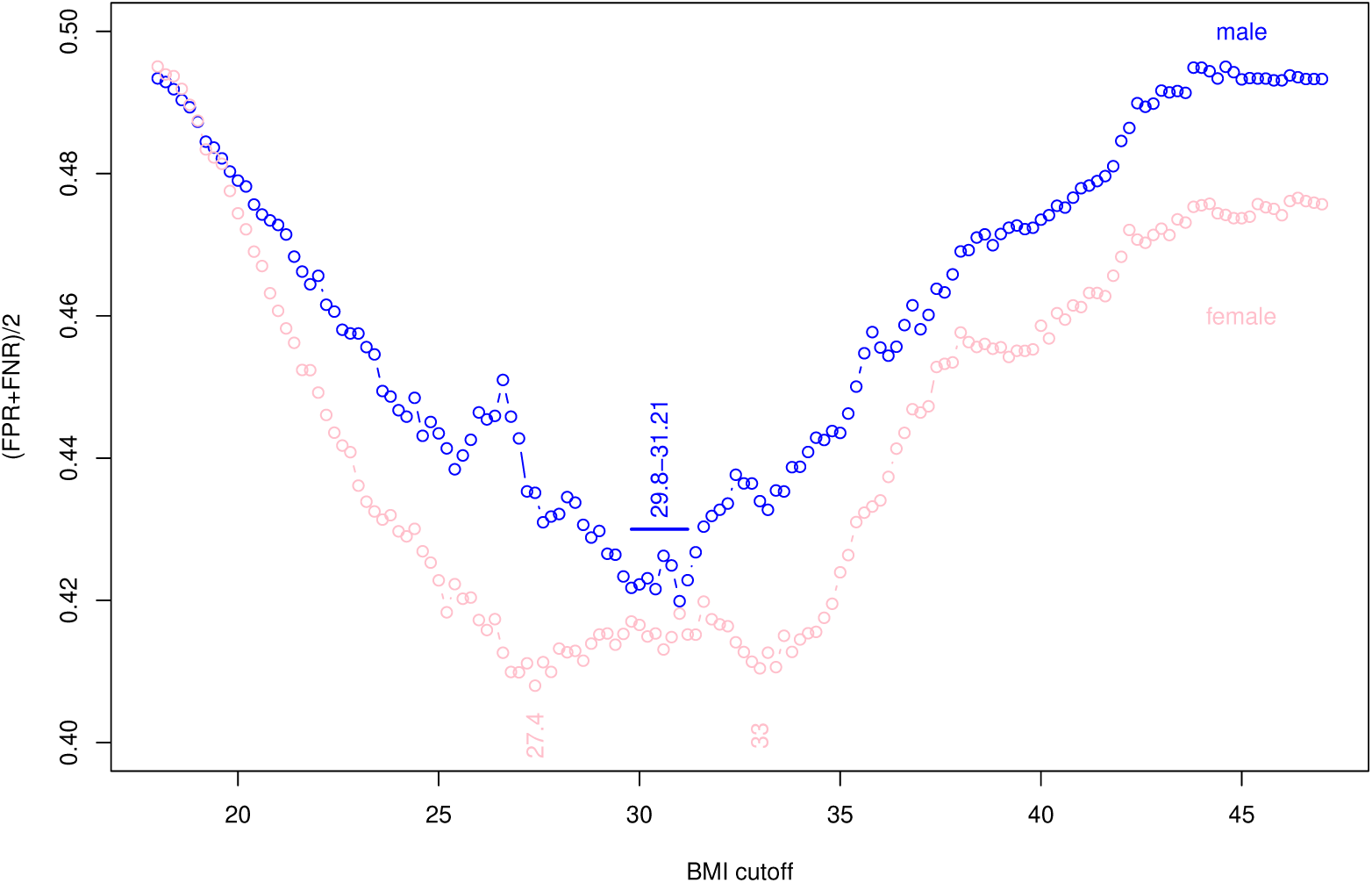
The mean of false positive and false negative rates as a function of BMI cutoff point (diabetes as the outcome variable) for male (blue) and female (pink) samples separately.

### Interpretable random forests with more independent variables

We re-ran SIRUS without age and gender but with a new set of other co-variates: total cholesterol, pulse, (average of three) diastolic blood pressure, education level, income level, and ethnicity, as well as BMI and diabetes status. The number of samples with all these information is reduced from 5175 to 4242. We relax the condition in SIRUS to output 15 rules instead of 10 in default. Table 2 shows the rules produced by SIRUS that involve BMI.

**Table 2:** Rules produced from SIRUS with BMI, total cholesterol, pulse, average of three diastolic blood pressure measurements, educational level, income level, and ethnicity as independent variables, and diabetes as the dependent variable. We relax the condition in SIRUS to print 15 rules, and six reported rules involve BMI. Three rules are BMI only, two rules involve BMI and educational level, and one rule involves BMI and total cholesterol. Similar to Table 1, FNR, FOR, FPR, FDR, (FPR+FNR)/2, (FOR+FDR)/2, odds-ratio, and Fisher test *p*-value are calculated.

| condition0 | $n_1$ | FNR | FOR | condition1 | $n_2$ | FPR | FDR | $\frac{FPR+FNR}{2}$ | $\frac{FOR+FDR}{2}$ | pv | OR |
| --- | --- | --- | --- | --- | --- | --- | --- | --- | --- | --- | --- |
| BMI < 25.6 | 1257 | 0.185 | 0.118 | BMI >25.6 | 2985 | 0.678 | 0.781 | 0.431 | 0.45 | 2E-15 | 2.1 |
| BMI < 30.4 | 2534 | 0.471 | 0.149 | BMI >30.4 | 1708 | 0.373 | 0.752 | 0.422 | 0.451 | 1.3E-15 | 1.88 |
| BMI < 34.9 | 3381 | 0.696 | 0.165 | BMI >34.9 | 861 | 0.179 | 0.715 | 0.437 | 0.44 | 1.9E-14 | 2 |
| BMI < 30.4<br>and edu ≥ HS | 2343 | 0.403 | 0.138 | else | 1899 | 0.413 | 0.748 | 0.408 | 0.443 | 3.8E-21 | 2.1 |
| BMI < 34.9<br>& edu ≥ HS | 3127 | 0.604 | 0.155 | else | 1115 | 0.231 | 0.715 | 0.418 | 0.435 | 3.4E-20 | 2.17 |
| BMI < 34.9<br>& chol ≥ 165 | 2396 | 0.394 | 0.132 | else | 1846 | 0.395 | 0.737 | 0.395 | 0.435 | 7.2E-27 | 2.35 |

The three rules, only involving BMI in Table 2, with cutoff values at 25.6, 30.4, 34.9, are similar to those in Table 1. The reason that these are slightly different is because the samples used are slightly different.

The next two cutoff values for BMI involve the education label: if BMI is lower and educational level is at least high school, the risk for diabetes is lower. Comparing the BMI only rules without considering the educational level, the new rules have lower FNR, FOR, FDR (but higher FPR). The Fisher’s test *p*-values are even smaller, not only because the sample sizes in the two nodes are more balanced, but also the odds-ratio is larger. Adding the education level in the consideration of BMI cutoff value enhances the signal.

The last rule in Table 2 involves, besides BMI, total cholesterol: samples with BMI *<* 34.9 and total cholesterol ≥ 165 have lower risk for diabetes. This new rule achieves the best *p*-value and the best odds-ratio, lower FNR, lower FOR, than the similar rule without considering cholesterol, though higher FPR and higher FDR.

In order to check the impact of total cholesterol on BMI cutoff directly, Fig. 5 plots the (FPR+FNR)/2 as a function of BMI cutoff for two subsets stratified by the total cholesterol level: those with cholesterol *<* 165 (in blue) and those with cholesterol ≥ 165 (in red). The determined BMI cutoff (around 25.4) for the low cholesterol group is much lower than that (BMI=30.8 or in the region of BMI=(29.8-34)) for the higher cholesterol group. The last rule in Table 2 (i.e., BMI *<* 34.9 and cholesterol ≥165 subset has lower risk of diabetes) is a reflection of the fact seen in Fig. 5 that the BMI cutoff for individuals with elevated cholesterol is higher.

**Figure 5:**
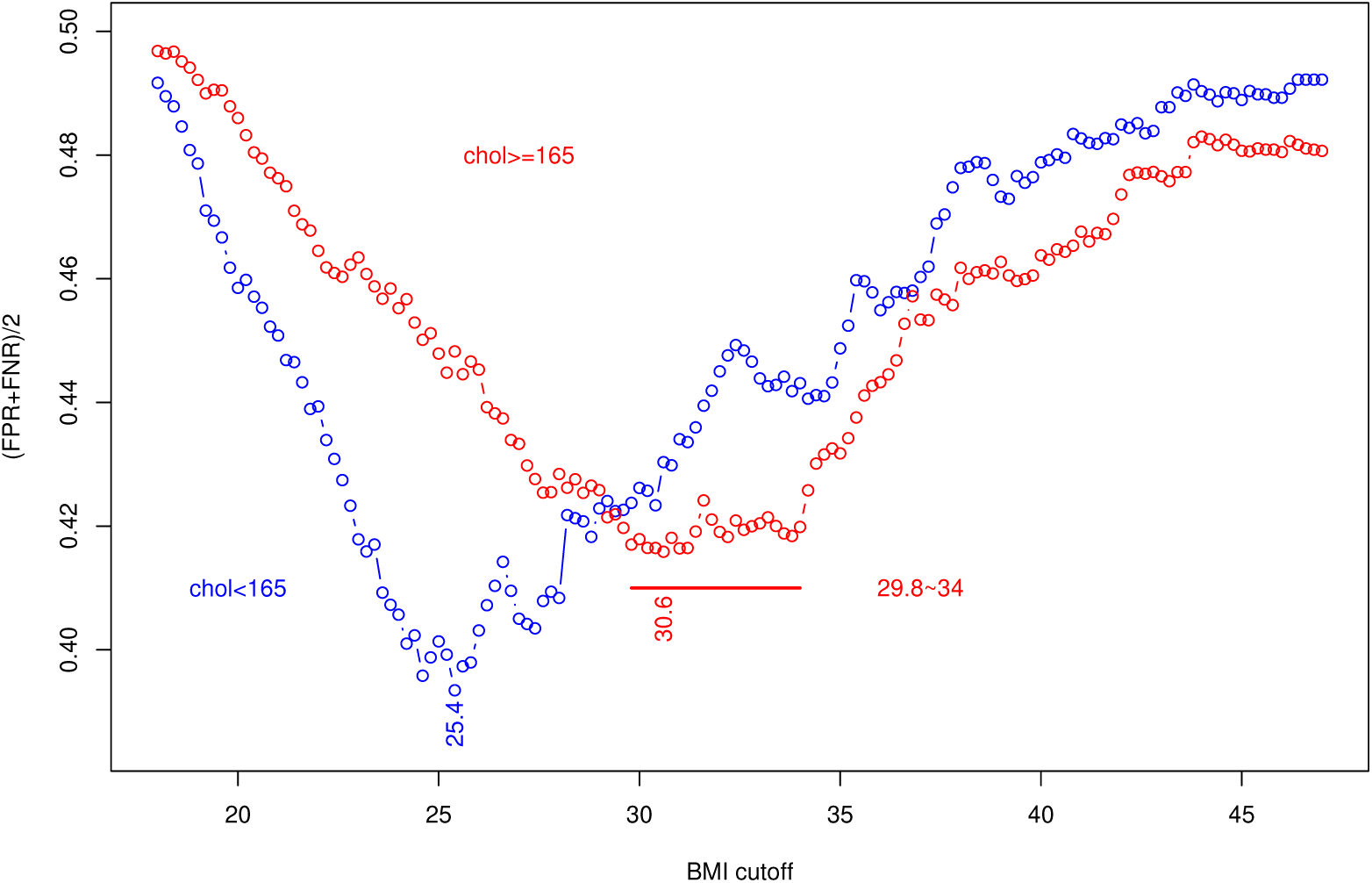
The mean of false positive and false negative rates as a function of BMI cutoff point (diabetes as the outcome variable) for high-cholesterol (red) and low-cholesterol (blue) group separately (diabetes as the outcome variable).

In both Table 1 and 2, the composite rules (“BMI ≥ 30.3 and male”, “BMI *<* 34.9 and cholesterol ≥165”) do not actually produce a new BMI cutoff value. However, these rules pick one BMI cutoff out of three choices (23.9, 30.3, 35 in Table 1, and 25.6, 30.4, 34.9 in Table 2) according to context provided by another variable (gender in Table 1 and cholesterol in Table 2). These choices are consistent with Fig. 4 and Fig. 5, in that for male gender the best BMI cutoff is around 30, where for people with high cholesterol the best BMI cutoff is around 35.

## Discussion

It was mentioned in the Introduction section that doctor recommended cutoff points between normal and abnormal clinical measures are not based on population distribution. Fig.6 shows the histogram of BMI for the n=5175 samples from NHANES used here. The mean bracketed by one standard deviation, median bracketed by one MAD (median of absolute deviance (Leys et al., 2013)), and mean bracketed by one standard deviation in the log-scale, are shown at the top. We also reproduced the estimated range of possible cutoff points from Figs.1-5 in Fig.6. It can be seen that all these cutoff points are within one standard deviation (or one MAD, or one one standard deviation in log scale) from the mean (or median). In other words, most people with doctor-claimed abnormal BMI values are not outliers, defined as those a few standard deviations away from the mean (Cousineau and Chartier, 2010) This is consistent with the statement “BMI percentiles may not adequately define risk of comorbid conditions” (Daniels, 2009).

**Figure 6:**
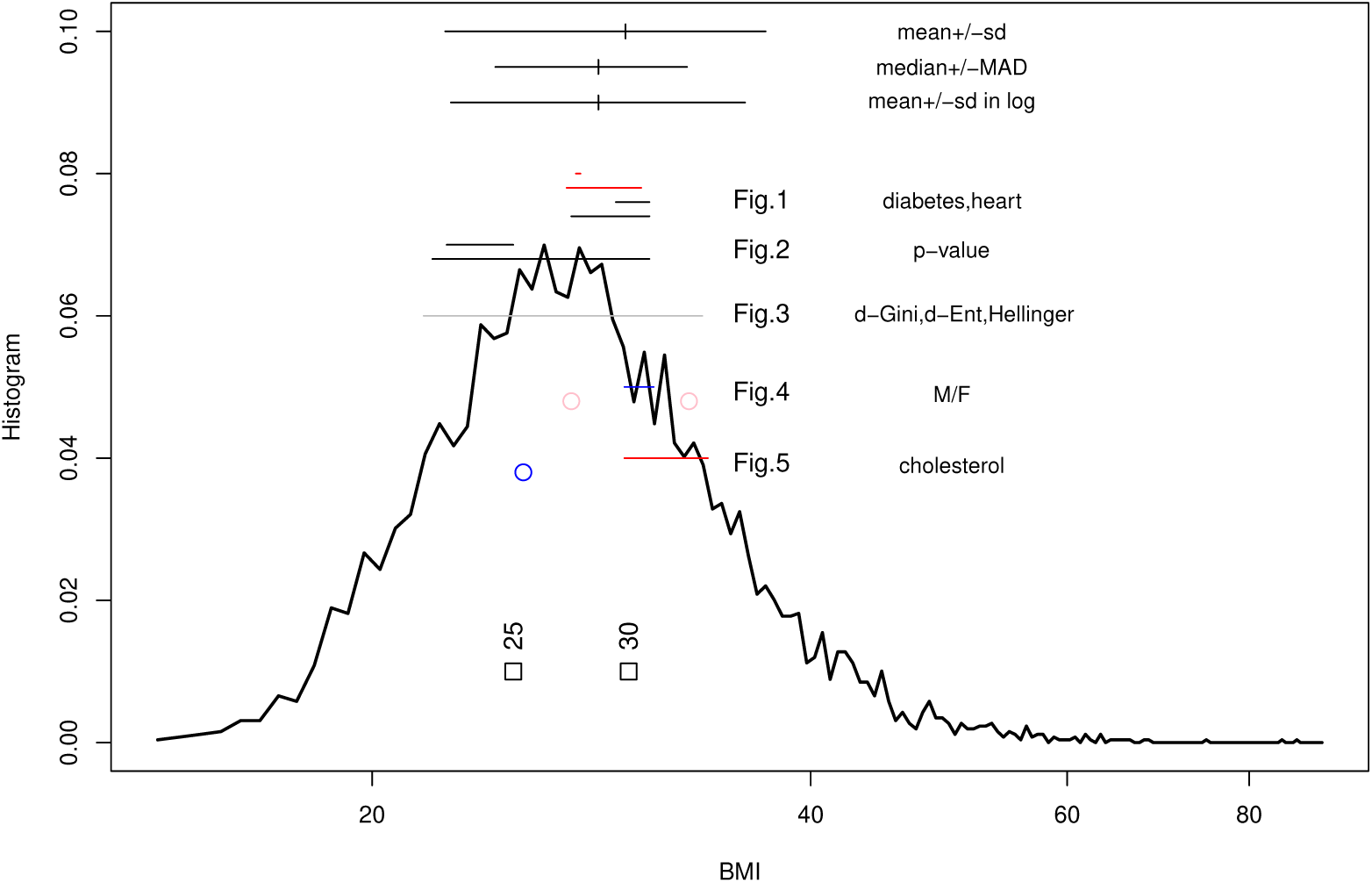
Histogram of BMI (in log-scale). The official cutoff points for overweight (BMI=25) and for obesity (BMI=30) are marked at the bottom. The population mean or median plus and minus one standard deviation (both normal and log scale) or one MAD (median of absolute deviance) are shown at the top. Other ranges of possible cutoff points from Figs.1-5 are reproduced (for more details, see Figs.1-5).

The usefulness of BMI has long been debated. A recent commission report distinguishes obesity (excessive adiposity) from clinical obesity, with the latter defined as “chronic, systemic illness characterized by alterations in the function of tissues, or organs, or the whole body, due to excessive adiposity” (Rubino et al., 2025). If BMI, as well as hip or waist size, is a reasonable measure of obesity, it may not lead, or has not yet led, to the clinical obesity. The report admits that obesity-associated diseases “rise as a continuum” with increasing levels of fat mass, and a binary label (preclinical and clinical obesity, or health and illness) is only “for clinical and policy-related purpose”. These statements imply that the cutoff between the two labels is arbitrary. Another statement made in in (Rubino et al., 2025) is “BMI should be used only as a surrogate measure of health risk at a population level, for epidemiological studies, or for screening purposes, rather than as an individual measure of health”.

Both highlighted conclusions from (Rubino et al., 2025) are consistent with the points made in our paper, that of the artificial nature of cutoff in a continuous risk function for individuals, and the cutoff being a balance point at the population level between two false classification rates. The balance has to be at the population level because for an individual person, false classification rate is either 0% or 100%. Because population is important, the cutoff determination will depend on the dataset used, on the medical outcome chosen, and on the cost of objective function. If these factors are fixed, in principle, the cutoff point is precisely defined. On the other hand, an unavoidable corollary is that when data, or medical outcome of concern, or cost and objective function, are different, the resulting cutoff point may also change. Further works are needed to check if our definition of cutoff points match those by medical experts, beyond BMI to any health-related measurement.

We believe that interpretable random forest is ideally suitable for our task of cutoff point determination. Although there are more sophisticated machine learning methods, such as artificial neural networks (ANN) and deep learning which is a subset of ANN, making them interpretable is not easy (Angelov and Soares, 2020; Antamis et al., 2024). The essence of making an algorithmic output interpretable is to have a “sparse” representation (Rudin, 2019) that can be trusted (Riberio et al., 2016) for making decision, as well as close to the actual mechanism. Although many commonly applied machine learning techniques are somewhat similar for classification tasks (Musolf et al., 2022), random forest and decision trees most resemble our goal of cutoff point determination. Simple rules are extracted from interpretable random forest based on how frequent they appear in node splitting of individual decision trees. Single-variable cutoff determination problem has been extensively studied, using a variety of quantities taken from the ROC (receiving operating characteristic) (Perkins and Schisterman, 2006; Rota and Antolini, 2014; Santos et al., 2019; Wang and Tian, 2024; Hassanzad and Hajian-Tilaki, 2024), such as Youden index (Youden, 1950) being equal to 1-FDR-FNR, (0,1)-distance (or Northwest distance) which is 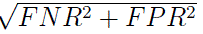, product method (or Southeast area) (Liu, 2012) which is (1-FNR)(1-FPR), and others (Unal, 2017). These choices are all some combinations of FPR and FNR, and are usually comparable to each other. The method proposed in this article is fundamentally different in that it is not single-variable based, thus belonging to a very different category.

Since the fundamental data structure in the single-variable cutoff problem is a 2-by-2 count table, it is natural to consider other measures beyond FNR and FPR. The first choices, from the common practices in other fields, such as genetic case-control association studies (Li, 2007) are (diagnostic) odds-ratio (DOR) (Glas et al., 2003), *χ*^2^ statistics (Miller and Siegmund, 1982), and (chi-square or Fisher) test *p*-values. However, it was pointed out that DOR tends to reach maximum at the borders instead of interior of the *x* range (Böhning et al., 2010; Hajian-Tilaki, 2017), and *p*-value only measures deviation from an unrealistic null hypothesis (Rota and Antolini, 2014). Our result does show that *p*-value as a cost function may lead to a different minimal, but it seems to still point to an overall correct region.

We have shown whole spectra of cost or object functions in Figs.1-5 instead of just reporting the minimum or maximum position. These plots show that there could be a region where the cost (objective) function values are low (high), indicating that the precise cutoff point is not as important as one might think, even though we can determine that point precisely. If a precise normal range of medical measures is less important, it can have consequence to general practitioners.

The application of interpretable random forest also addresses other issues, such as its ability for variable selection (i.e., which node-splitting variable is included in the rule summary), continuous or ordinal medical outcomes (change *y* from binary variable to a continuous variable), when to stop the tree branching (reminiscent of the stopping segmentation in DNA sequences (Li, 2001) and other change-point analysis in time series), etc. These issues will be addressed in future studies.

## Data and Programs

### Population data

The NHANES (National Health and Nutrition Examination Survey) data from 2017-2018 (https://wwwn.cdc.gov/nchs/nhanes/continuousnhanes/default.aspx?BeginYear=2017) is used, with these files being downloaded: DEMO J (demographic variables and sample weights), BMX J (body measures), BPX J (blood pressure),. TCHOL J (cholesteroltotal), DIQ J (diabetes), MCQ J (medical conditions).

The main outcome variable is DIQ010 (“doctor told you have diabetes”). Another medical condition used in Fig.1 is a logic OR operation on five heart problems: MCQ160B (“ever told had congestive heart failure”), MCD160C (“ever told you had coronary heart disease”), MCD160D (“ever told you had angina pectoris”), MCD160E (“ever told you had heart attack”), MCD160F (“ever told you had a stroke”).

The main independent variable whose cutoff is examined is BMXBMI (body mass index). Other covariates used include: BPXPLS (pulse), BPXDI1,BPXDI2,BPXDI3 (distolic blood pressure measures 1,2,3), BPXSY1,BPXSY2,BPXSY3 (systolic blood pressure measures 1,2,3), LBXTC (total cholesterol mg/mL), DMDEDUC2 (education level, 1/2/3/4/5 for less than 9th grade, 9-11 grades, high-school, college, college graduate), INDHHIN2 (income level), RIDRETH3 (race, 1/2/3/4/6/7 for Mexican/Hispanic/White/Black/Asian/Mixed). The age and gender variable are RIDAGEYR and RIAGENDR (1/2 for male/female).

The data is filtered for adult (age equal or older than 20) samples only (n=9254-3685 = 5569). Of these, n=5175 have BMI information,

### Programs used

All calculation was carried in the R environment (https://www.r-project.org/).

The reading of the NHANES SAS format files (XPT) was carried out by the *SASxport* program (https://github.com/r-gregmisc/SASxport), and data manipulation by *nhanesA* program (https://github.com/cjendres1/nhanes) (Ale et al., 2024). The program used for interpretable random forests is the SIRUS program (https://gitlab.com/drti/sirus) (Bénard et al., 2021).

## Data Availability

Data is not generated by the author, but downloaded from public domain

https://www.cdc.gov/nchs/nhanes/index.html

## Acknowledgement

I would like to thank Yaning Yang, Yannis Almirantis, Weihao Wang, Xiancheng Yang, and Fengguo Sun for discussions and suggestions.

## Appendices

### A.1 The four groups of samples organized into 2-by-2 count table

If an increase of the value of *x* also increases the probability of *y* = 1 (risk), we use a cutoff *X* for *x* to partition the samples into high-risk group if *x > X*, and low-risk group if *x < X*. The true value of *y* is either 0 (healthy group) or 1 (disease group). All samples can be separated into four groups: true negative TN (*x < X* and y=0), false negative FN (*x < X* and y=1), false positive FP (*x > X* and y=0), and true positive TP (*x > X* and y=1), forming a 2-by-2 matrix (see Table A.1).

The off-diagonal elements of the matrix in Table A.1 are false calls (FN and FP). There are two different ways to normalized these false counts to obtain error rates. One is by normalizing the false counts by the actual/true number of *y* = 0 and *y* = 1 samples, leading to false positive or negative rates (FPR or FNR): *P* (*x > X*|*y* = 0) or *P* (*x < X*|*y* = 1). There are other names for FPR and FNR (or 1-FPR and 1-FNR), such as specificity(=1-FPR), sensitivity or recall (=1-FNR). We do not use other names in this paper to avoid confusion.

Another way to normalize the false counts is to divide them by the number of samples by the cutoff partition, leading to false discovery rate (FDR) and false omission rate (FOR): *P* (*y* = 0|*x > X*)*andP* (*y* = 1|*x < X*). Comparing the two ways of normalizing the false counts, FPR and FNR are closely tied to hypothesis testing framework where FPR is type-I error rate, and FNR is the type-II error rate (and 1-FNR is the statistical power); whereas FDR and FOR are often the printed error rates in data analysis programs such as decision trees and random forests.

### A.2 Cutoff determination with two normal distributions for two medical outcomes

Suppose the distribution of a measurement *x*, in samples both with a medical outcome (*y* = 1 or plus, e.g. with a disease) or not (*y* = 0 or minus, e.g. being healthy), follow a normal distribution, but with different means (e.g. *µ*_+_ = 10 for *y* = 1, and, *µ_−_* = 5 for *y* = 0), though same variance (*σ*_+_ = *σ_−_* = 1); the population prevalence/proportion of those with the medical outcome *P*_+_ is assumed to be (e.g) 0.05. The probability of the medical outcome given the *x* value can be determined by the Bayes’ theorem:

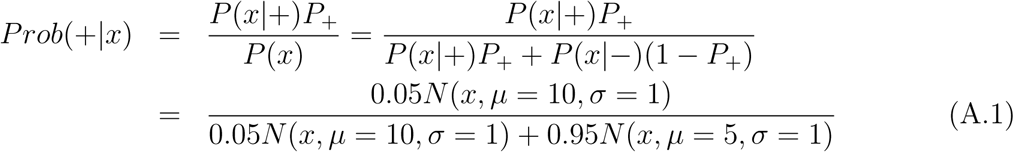

**Table A.1:**
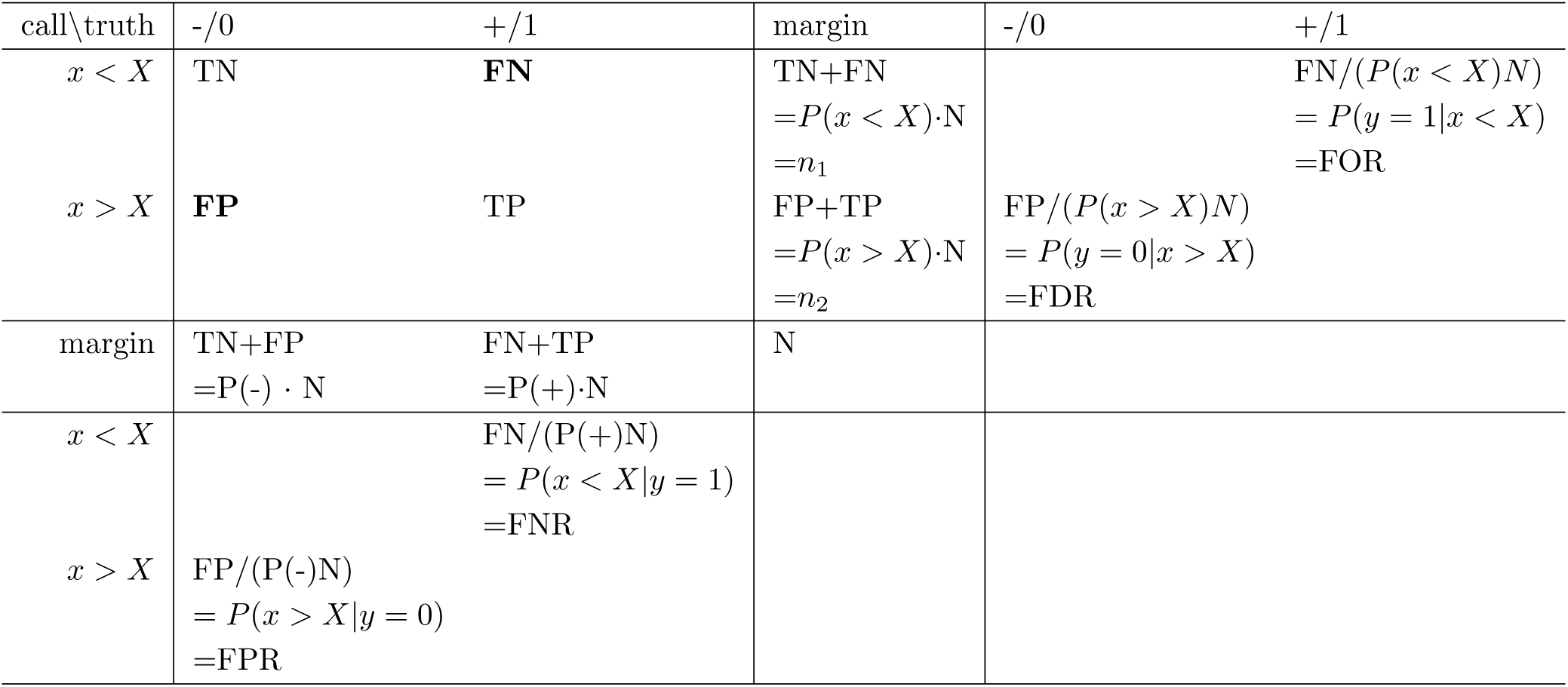
Illustration of the notations used in this paper: (1) number of samples in the four groups when a cutoff point *X* is set: TN (true negative), FN (false negative), FP (false positive), TP (true positive); (2) false rates by normalizing numbers of positives (unhealthy) and negatives (healthy): FNR (false negative rate), FPR (false positive rate); (3) false rates by normalizing number of samples larger or smaller than the cutoff value: FDR (false discover rate), FOR (false omission rate).

Fig. A.1(A) shows *P* (*x*|+), *P* (*x*|−), *P* (*x*), and *P* (+|*x*). *P* (+|*x*) is a continuous function of *x* without a discontinuous jump or “threshold effect”. This conclusion is unrelated with the prevalence value chose: another *P* (+|*x*) curve is plotted in Fig.A.1(A) with P(+)=0.5 (pink), which remains to be continuous.

If one uses the *x* value that minimizes the undesirable medical outcome as the cutoff point, the value would be as low as possible, all the way to *x* = 0. If one chooses the *x* corresponding to 5% to have a bad outcome (i.e., *P* (*y* = 1|*x*) = 0.05) as the cutoff point, that value depends on our particular choice of 5%, which is arbitrary, and therefore is also arbitrary.

The arbitrariness problem can be solved, however, if we choose the *x* value that minimizes the cost function (FPR+FNR)/2. The reason for the existence of a unique point is the following: FPR always decreases with increasing cutoff level, whereas FNR always increases; therefore, there will be middle *x* value where the two types of errors are balanced (see Fig.A.1(C), at *x* = 7.5).

When the FPR is heavily weighted (*w*_0_ =0.95, equal to *P* (*y* = 0)), it will dictate the trend of the cost function which will, for most of the *x* range, decrease with higher cutoff values. However, the less weighted FNR (*w*_1_ = 0.05, still plays a role, which will eventually help the cost function to reach a balance (*x* = 8.82, Fig.A.1(C)). This minimum point can be reached in a theoretical model because the two normal distributions will have infinitely tails. For real data, both histograms will have limit, then a balance between the heavy false positive and the light false negative may not be reached.

From Fig.A.1(A), *x* = 7.5 is also the point where *P* (*x*|−) = *P* (*x*|+). i.e., where the two distributions, *P* (*x*|−) and *P* (*x*|+), intersect. Because *P* (*x*|−) and *P* (*x*|+) have the same shape in our model, the tail area of *P* (*x*|−) at this cutoff point, which is FPR, and the head area of *P* (*x*|+), which is FNR, are also the same. Using FNR=FPR, or equivalently, sensitivity=specificity, as a criterion to determine the cutoff point, is called *ϕ* index method in (Santos et al., 2019).

One may curious about using the mean of false discover rate and false omission rate, (FDR+FOR)/2, as the cost function for cutoff point determination. However, both FDR and FOR contain population prevalence information (see Table A.1, where denominator for, e.g., FDR, FP+TP, is affected by the prevalence). This could make the cost function mostly dominated by one error rate, as seen in Fig.A.1(D), where the minimal is reached at *x* = 10.5. Since the two errors do not seem to be easily balanced, (FDR+FOR)/2 may not be a good choice for the cost function.

The difference between the two cumulative distributions, *F* (*x*|−) − *F* (*x*|+), can be used for cutoff determination (Hsieh and Turnbull, 1996). The cutoff point is where the *F* (*x*|−) − *F* (*x*|+) reaches the maximum, because that’s also the maximum point for sum of sensitivity and specificity, or minimum point for sum of FNR and FPR. Fig.A.1(B) shows the cumulative distributions for healthy and diseased group, *F* (*x*|−) (in black) and *F* (*x*|+) (in blue), as well as their difference (in red). The maximum of the latter is reached at *x* = 7.5.

**Figure A.1:**
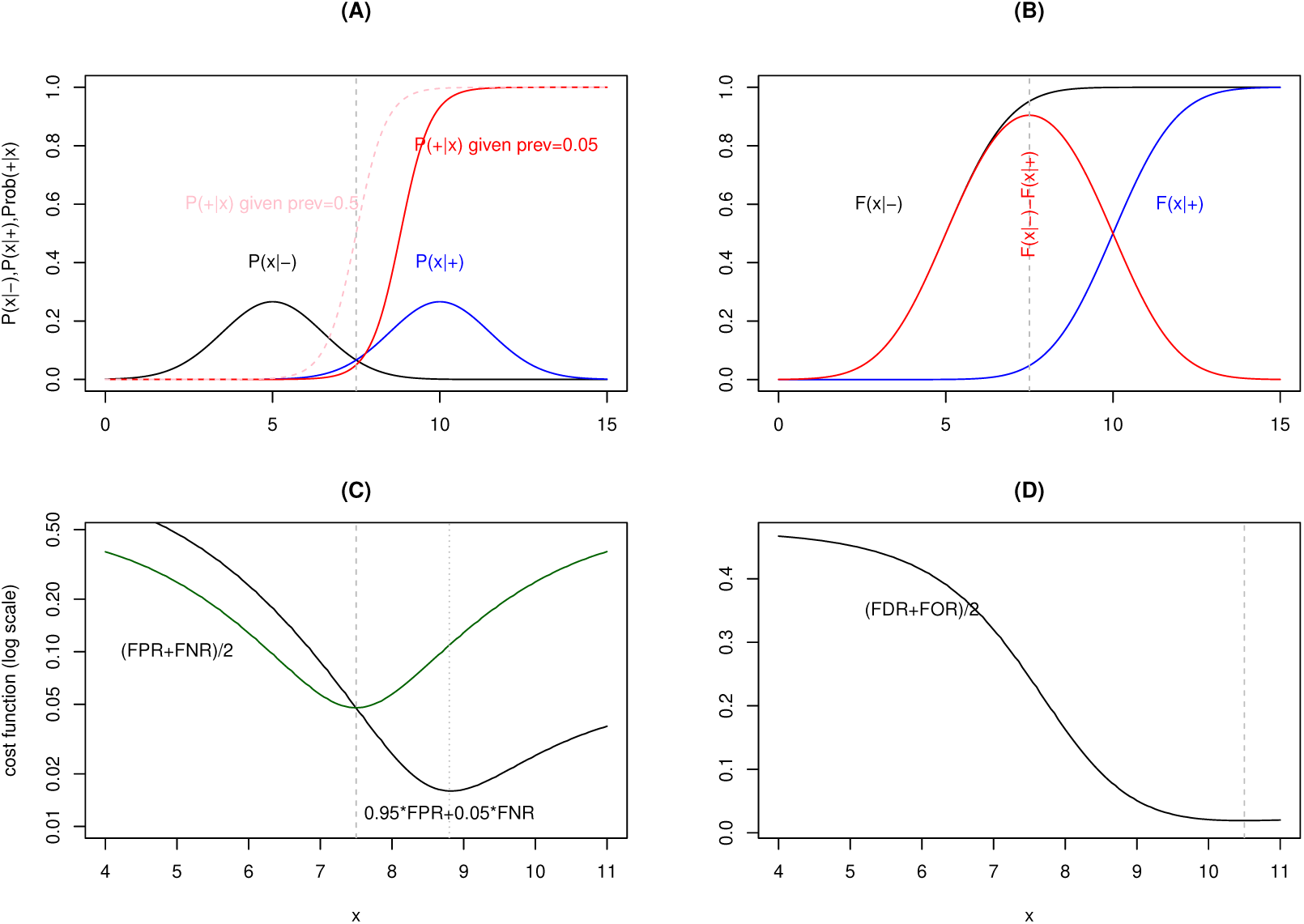
Illustration of a theoretical model where the diseased group (+) and healthy group (−) has distinct distribution of independent variable *x*: *N* (10, 1) (normal distribution with mean of 10, standard deviation of 1) for the positive group, *N* (5, 1) for the negative group. (A) probability density function (pdf) for healthy and diseased group, *P* (*x*|−) (black), *P* (*x*|+) (blue), and the probability (risk) for disease assuming the disease prevalence to be 0.05 (red) or 0.5 (pink). (B) The cumulative density function (cdf) for the healthy and the diseased groups, *F* (*x*|−) (black), *F* (*x*|+) (blue), and their difference *F* (*x*|−) − *F* (*x*|+) (red). The maximum of the difference is reached at *x* = 7.5. (C) The mean of false positive and false negative rates ((FPR+FNR)/2), and a weighted average ( 0.95 FPR+0.05 FNR) of the two rates, as a function of *x*. (D) The mean of false discover rate and false omission rate, (FDR+FOR)/2, as a function of *x*.

